# Global, regional, and national estimates of target population sizes for COVID-19 vaccination

**DOI:** 10.1101/2020.09.29.20200469

**Authors:** Wei Wang, Qianhui Wu, Juan Yang, Kaige Dong, Xinghui Chen, Xufang Bai, Xinhua Chen, Zhiyuan Chen, Cécile Viboud, Marco Ajelli, Hongjie Yu

**Affiliations:** School of Public Health, Fudan University, Key Laboratory of Public Health Safety, Ministry of Education, Shanghai, China; Division of International Epidemiology and Population Studies, Fogarty International Center, National Institutes of Health, Bethesda, MD, USA; Department of Epidemiology and Biostatistics, Indiana University School of Public Health, Bloomington, IN, USA; Laboratory for the Modeling of Biological and Socio-technical Systems, Northeastern University, Boston, MA USA

**Author notes:** **Corresponding Authors:** Hongjie Yu, PhD, School of Public Health, Fudan University, Key Laboratory of Public Health Safety, Ministry of Education, No. 138 Yixueyuan Road, Xuhui, District, 200032, Shanghai, China. These authors are joint senior authors and contributed equally to this work (Cécile Viboud, Marco Ajelli, Hongjie Yu).

**Keywords:** target population, heterogeneous distribution, vaccination, coronavirus

## Abstract

**Background:** COVID-19 vaccine prioritization and allocation strategies that maximize health benefit through efficient use of limited resources are urgently needed. We aimed to provide global, regional, and national estimates of target population sizes for COVID-19 vaccination to inform country-specific immunization strategies on a global scale.

**Methods:** Based on a previous study of international allocation for pandemic COVID-19 vaccines, we classified the entire world population into eleven priority groups. Information on priority groups was derived from a multi-pronged search of official websites, media sources and academic journal articles. The sizes of different priority groups were projected for 194 countries globally.

**Results:** Overall, the size of COVID-19 vaccine recipient population varied markedly by goals of the vaccination program and geography. The general population aged <60 years without any underlying condition accounts for the majority of the total population (5.2 billion people, 68%), followed by 2.3 billion individuals at risk of severe disease, and 246.9 million essential workers which are critical to maintaining a functional society. Differences in the demographic structure, presence of underlying conditions, and number of essential workers led to highly variable estimates of target populations both at the WHO region and country level. In particular, Europe has the highest share of essential workers (6.8%) and the highest share of individuals with underlying conditions (37.8%), two priority categories to maintain societal functions and reduce severe burden. In contrast, Africa has the highest share of healthy adults, school-age individuals, and infants (77.6%), which are the key groups to target to reduce community transmission.

**Interpretation:** The sizeable distribution of target groups on a country and regional bases underlines the importance of equitable and efficient vaccine prioritization and allocation globally. The direct and indirect benefits of COVID-19 vaccination should be balanced by considering local differences in demography and health.

## Introduction

As coronavirus disease 2019 (COVID-19) continues to spread across the world, more than 200 candidate vaccines for COVID-19 are in development and 9 candidates have entered in phase III clinical trials as of September 19, 2020 (*1*). Hopes are high to bring one or more vaccine candidates to market by the end of the year. Despite 30.3 million cases reported so far (*2*), most of the world population still remain susceptible – an increasing number of sero-epidemiological studies are finding low seroprevalence of antibodies to SARS-CoV-2, in the range 1.0%-10.8% (*3-5*), although higher incidences were reported locally [e.g., ∼20% in New York City, NY, USA (*6*)]. As such, a large demand for COVID-19 vaccine is expected in the next year.

An internal survey of 37 members of the Developing Countries Vaccine Manufacturers has revealed that the global production capacity is estimated to be ∼3.5 billion doses annually (*7*). Thus, given a two-dose vaccination schedule which is planned for the majority of current COVID-19 candidates, current annual production capacity (*7*) will be too limited to achieve herd immunity by immunizing 60%-80% of the global population. As such, defining a prioritized vaccination program will be necessary.

There are important questions about equitable and efficient distribution of COVID-19 vaccine as many low- and middle- income countries lack COVID-19 vaccine research, development and production (*8*). To bring the pandemic under control via equitable access to COVID-19 vaccines, COVAX, the vaccine pillar of the Access to COVID-19 Tools (ACT) Accelerator, has been established with global cooperation to ensure availability to both higher-income and lower-income countries (*9*). In addition, given the likelihood of an initial period of vaccine shortage, country-specific interim frameworks for COVID-19 vaccine allocation and distribution have been developed by experts in the United States (*8, 10*) and United Kingdom (*11*). However, information is lacking about the number of vaccine doses that each region and country needs. This will hamper the equitable and efficient allocation and distribution of COVID-19 vaccine.

Here, we provide global, regional, and national estimates of the size of the COVID- 19 vaccine recipient population by priority group under the allocation frameworks proposed by various international teams (*8, 10-12*). The vulnerability of each country to COVID-19 is based upon factors such as geographical location, disease burden, the likelihood of an outbreak and the potential for subsequent severe public health impacts. Priority groups can be categorized into different allocation tiers according to country-specific pandemic characteristics and vaccine objectives. Estimates of target population sizes can guide relevant stakeholders in the development of fair and equitable global allocation strategies and inform vaccination programmes tailored to the local specificities of each population.

## Methods

### Definition of target populations for COVID-19 vaccination

Previous proposals for the international allocation for pandemic COVID-19 vaccines have endorsed three fundamental objectives (*8, 10-12*):

1. maintaining essential core societal functions during the COVID-19 pandemic, such as essential health services and food delivery;
2. protecting people from irreversible and devastating harm, such as death and severe COVID-19 disease that causes long-term organ damage (e.g., lung, kidney and liver);
3. controlling community transmission, enabling a return to normal pre-pandemic economic and social activities.

The importance of maintaining essential core societal functions has been highlighted in the context of COVID-19 pandemic, and the concept of essential workers has already been extended beyond health-care personnel (*13*). In light of previous proposals, these include, but are not limited to, workers in the food industry and domestic transportation, police and military staff who maintain public safety, as well as workers maintaining electricity, water, fuel, information, and financial infrastructures.

Regarding individuals who may experience irreversible and devastating harm from COVID-19, previous reports have identified those older than 65 years of age, those with high-risk health conditions, and those in close contact with people at very high risk of poor outcomes (e.g., nursing home and long-term care facility workers) as target population (*10, 12*).

A third possible vaccine goal is to reduce COVID-19 transmission; in this case, high transmission groups should be targeted. Target populations include adults and children involved in economic or educational activity, who experience higher risk of economic or educational harm from not working or going to school, and have a higher probability of transmission when going back to work or school due to increased contacts (*12*).

We defined groups of potential vaccine recipients aligned with the three goals of COVID-19 vaccination (Fig. 1). First, to maintain essential core societal services, individuals who are essential to maintaining an effective healthcare system (i.e., healthcare workers), national and social security (i.e., police and military), and normal living supplies (i.e., workers in essential infrastructures) need to be given careful consideration for priority.

**Figure 1.**
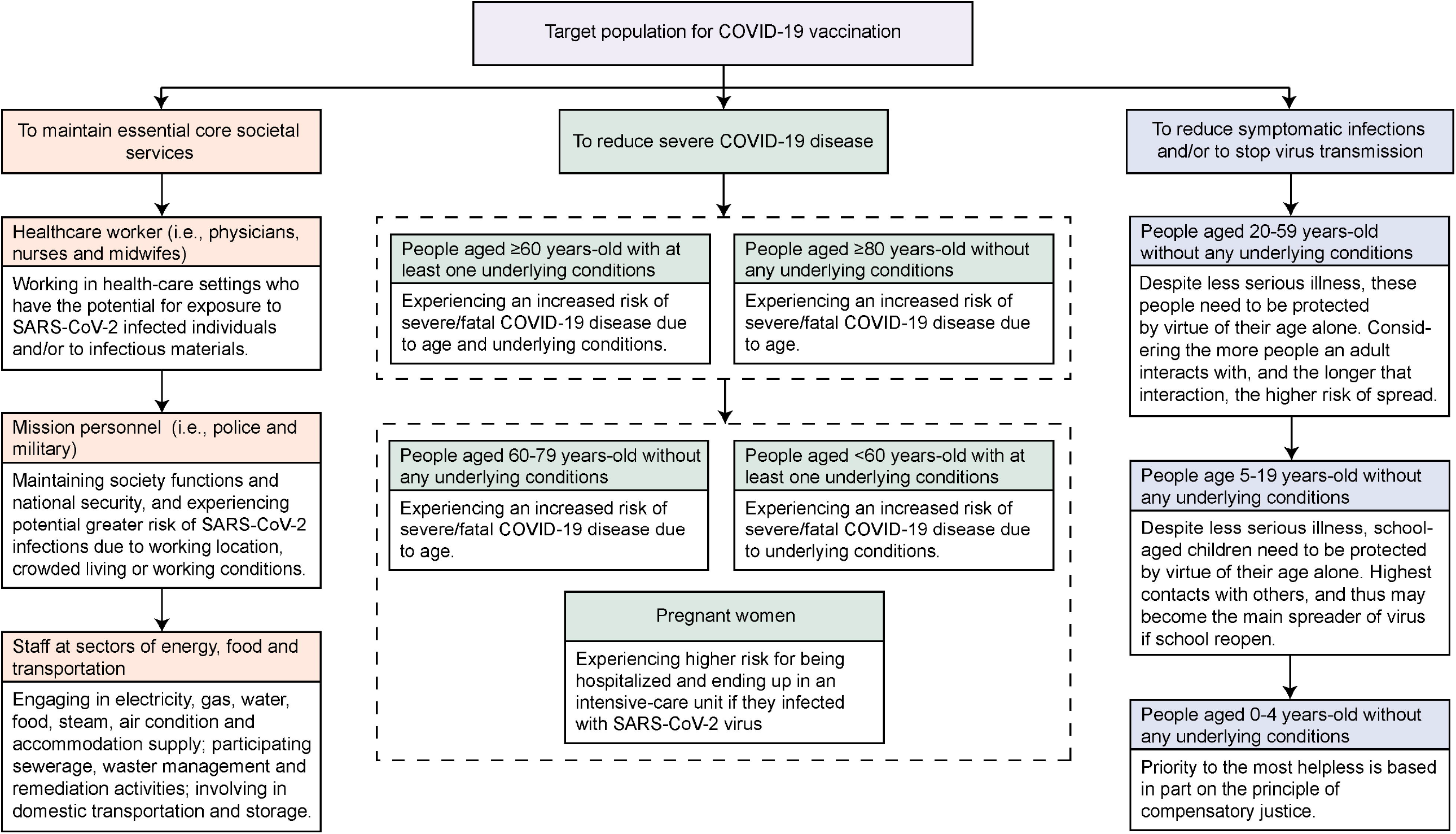
Priority group for COVID-19 vaccination.

Second, to reduce severe COVID-19 disease (i.e., hospitalizations, critical care admissions, and deaths), people with underlying conditions [i.e., cardiovascular disease, chronic kidney disease, chronic respiratory disease, chronic liver disease, diabetes, cancer with direct immunosuppression, cancer without direct immunosuppression but with possible immunosuppression caused by treatment, HIV/AIDS, tuberculosis (excluding latent infections), chronic neurological disorders and sickle cell disorders] (*14*), those older than 60 years of age without any underlying conditions, as well as pregnant women should be included as a candidate priority groups. Considering age-specific susceptibility to SARS-CoV-2 infection (*15*), we then reclassified people with and without underlying conditions into different groups by virtue of their age, i.e., people aged ≥60 or <60 years-old with at least one underlying condition, people aged ≥80 or 60-79 years-old without any underlying condition.

Third, to reduce symptomatic infections and/or to stop virus transmission, vaccination should extend to all individuals younger than 60 years of age without any underlying conditions. These individuals are further reclassified into three groups (i.e., people aged 20-59 years-old, those aged 5-19 years-old, and those aged 0-4 years-old) on the basis of their risk of transmitting virus and economic harm from not working (*15*).

### Data source

To estimate the size of priority groups for vaccination by country, we extracted information from publicly available data sources during 2013-2020 (see Tab. S1 for data sources), including the 1) United Nations (UN) mid-year population estimates for 2020 for 194 WHO member states (and countries/territories); 2) country-specific sizes of the military population from the World Bank Group or searching Baidu, Bing, and Google search engines using the search terms “military size” and World Health Organization country names; 3) the density of physicians, nurses and midwiferies by country from the World Bank and the World Health Organization; 4) the number of people working in the electricity, gas, water, steam and air conditioning sectors, food, accommodation, domestic transportation and storage industries, using census data on economically active population in 152 countries; 5) the number of individuals at increased risk of severe COVID-19 by age and country from previous report by Clark A, et al. (*14*). Two independent investigators applied the same search procedure for cross-checking and comprehensiveness.

### Multiple imputation of missing data

Up to 50 (25.8%) countries had missing values (see completeness analysis of data in Tab. S2) for the number of essential workers who ensure basic life needs. Thus, we employed a state-of-the-art Multivariate Imputation by Chained Equations” (MICE) algorithm (*16*) to impute missing values in the database. The number of multiple imputations was set as 5 with each imputation running 5 realizations. For each of the 5 realizations of imputed databases, we independently performed regression analysis of the size of each essential workers subgroup, using UN mid-year population, GDP per capita (current US$) and UN geographical regions as covariates, which are key determinants for the distribution of such populations. The final estimates were pooled from the 5 independent regressions on 5 imputed databases (i.e., mean predictive value).

### Estimates of the target population for COVID-19 vaccination

Global, regional, and national estimates of each target population were obtained by summing the relevant population group estimates stratified by vaccination goal (see data in Tab. S3). To avoid the overlap between the group of essential workers and adults aged 20-59 years without any underlying conditions, we subtracted those engaging in essential work activities from the broader group of healthy adults. Moreover, data on age-specific prevalence of underlying conditions were lacking for 11 countries. In the main analysis, we assume that the age-specific prevalence of underlying conditions in countries with missing data is the average of that in countries with available data in the same WHO region. Then, the number of persons with and without underlying conditions at a given age is equal to the prevalence of underlying conditions multiplied by the corresponding population size. In a sensitive analysis reported in Appendix, we assume that, when the data on underlying conditions is not available, the number of persons without any underlying condition corresponds the total number of persons of that age.

In main analysis, to consider vaccine programs tailored the epidemiological situation of individual countries, we also used COVID-19 case counts (as of September 13, 2020) and serology data to estimate the size of the population already infected, who may be at lower priority for vaccination. We found data on the number of laboratory-confirmed cases by RT-PCR (n=179 countries) or serological assays (n=15) from published literature and official reports. (see Tab. S4). The number of serologically-confirmed case in a country was measured as the seroprevalence of SARS-CoV-2 (*6*) multiplied by the corresponding population size. In sensitivity analyses, we excluded this epidemiological information from vaccine allocation estimates. In addition, to account for potential issues concerning vaccine hesitancy and delivery, a sensitive analysis was performed to estimate the size of COVID-19 vaccine recipient population by assuming a vaccination coverage of 60-80% (*17*).

## Results

### Global prospective

On a global scale, if a universal COVID-19 vaccination program was implemented, the target population would include 7.75 billion people (Tab. 1 and Fig. 2A). Of these, the target population that maintains core societal functions is estimated to be 246.9 million, including 40.7 million (16.5%) police and military personnel, 46.9 million (19.0%) healthcare workers, and 159.4 million (64.6%) individuals that maintain critical infrastructure and other important services, equal to 3.2% of the total population (Tab. 1). Among the 2.3 billion individuals at risk of severe COVID-19 disease who need to be vaccinated to minimize the health burden of COVID-19 (∼29% of the total population), 1.7 billion (∼22.4% of total population) individuals had at least one underlying condition, followed by those aged ≥60 years-old without any underlying conditions (0.4 billion, 4.9%) and pregnant women (0.1 billion, 1.8%). Considering an estimated annual vaccine production capacity of about 3.5 billion doses (*7*) and a (purely theoretical) homogeneous distribution of vaccines, vaccination may take at least 2 years for 2-dose schedules. The estimated size of the general population aged <60 years without underlying condition is estimated at 5.2 billion (∼68% of total population) (Tab. 1). If we subtract individuals who have experienced a natural SARS-CoV-2 infection and consider that they do not need to be vaccinated, the total target population decreases to 7.6 billion people (Tab. S4). The size of COVID-19 vaccine recipient population is estimated to be 4.7-6.2 billion people if vaccination coverage reaches 60-80% (Tab. S5) (*17*).

**Figure 2.**
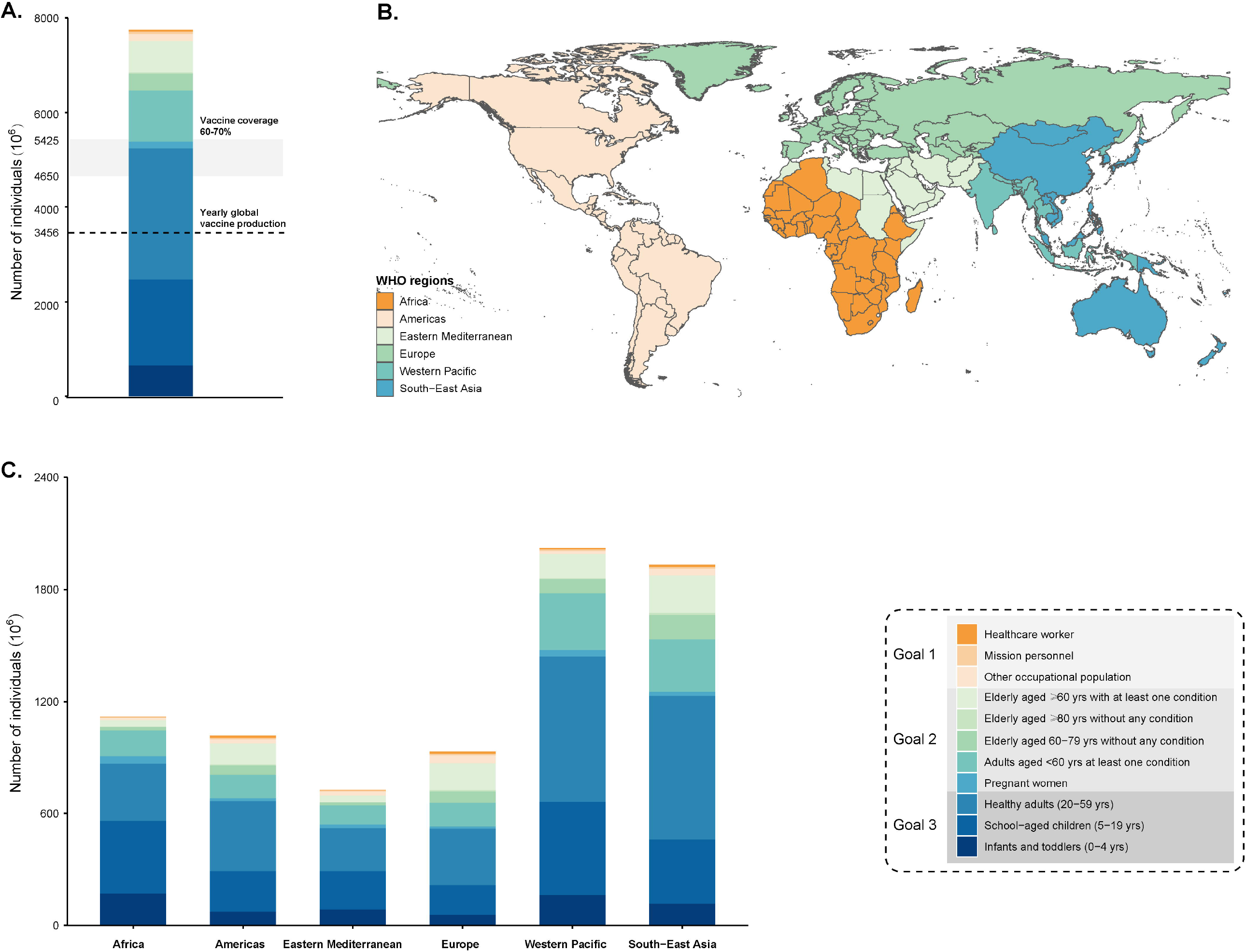
Global and regional estimates of the size of target populations for COVID-19 vaccination, by goals of vaccination program. (A) global estimates of the size of each target population for COVID-19 vaccination; (B) geographical distribution of member states by WHO regions; (C) regional estimates of the size of each target populations for COVID-19 vaccination.

### Regional perspectives

Geographical disparities are observed in the share of different target population groups across WHO regions. If we consider the entire population as target for vaccination (no prioritization by occupation or risk group), South-East Asia (2.0 billion, 26.1%) and Western Pacific (1.9 billion, 24.9%) together account for 51.0% of the population to vaccinate, while 14.5%, 13.1%, 12.0% and 9.4% of the target populations are distributed in Africa (1.1 billion), Americas (1.0 billion), Europe (0.9 billion) and Eastern Mediterranean (0.7 billion) regions, respectively (Tab. 1 and Fig. 2C).

In addition, the size of target population to maintain essential societal functions varies considerably by region, with highest concentration in Europe (63.8 million people, 25.8%), Western Pacific (58.8 million people, 23.8%), and the Americas (41.8 million people, 16.9%, Tab. 1 and Fig. 3). The size of the population at high risk of severe COVID-19 disease and those to vaccinate to contain COVID-19 are higher in Western Pacific (1.87 billion) and South-East Asia (1.99 billion).

**Figure 3.**
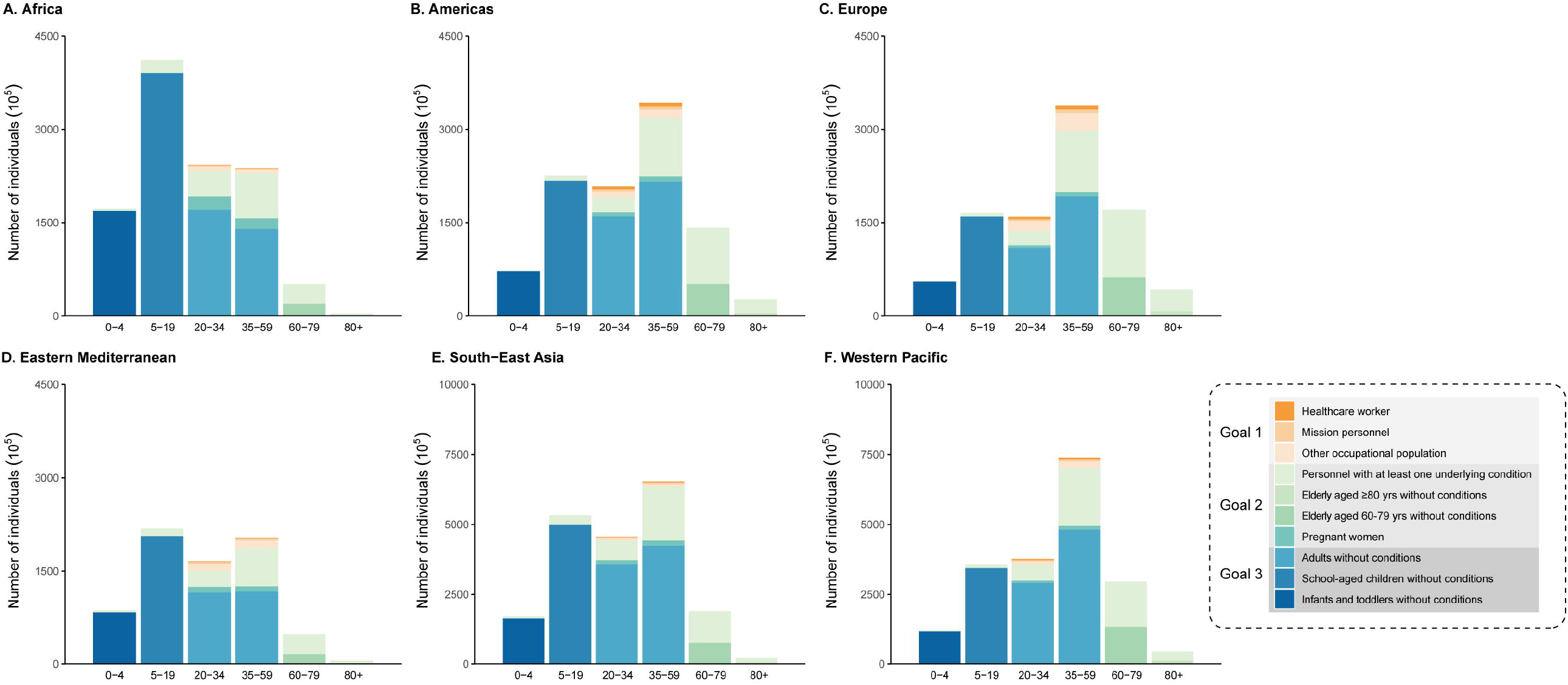
Age distribution of target populations for COVID-19 vaccination program across regions, by goals of vaccination program.

The size of each target population by age also varies markedly across regions. It is noteworthy that few individuals are over 80 years of age in Africa whilst a considerable fraction reside in Europe and North America (Fig. 3). The working-age population accounts for a substantially larger proportion of the total population than other target populations in all regions. In addition, the share of individuals aged <20 years of age is relatively high in Africa compared to other regions.

### National perspectives

On a country level, sizable heterogeneity emerges in the distribution of different target population groups, ranging from 1,000 people to 1.4 billion people (Fig. 4). National estimates of the size of target population suggest that seven countries, including China, India, United States, Indonesia, Pakistan, Brazil and Nigeria, have a larger share of total target population (Fig. 4); by contrast, countries in Africa and Eastern Mediterranean regions show a relatively lower share of total target population (Fig. 5 and Fig. S1). We also found that the target population that maintains essential core societal function is more predominant in middle- and high- income countries (Fig. 5 and Fig. S2). Moreover, between-country variations in the size of target population to reduce severe disease or to contain SARS-CoV-2 transmission were observed, with 51.2% of total population distributed in China, India, United States, Indonesia, Japan, Russian Federation and Brazil (Fig. 5, Fig. S3 and Fig. S4).

**Figure. 4.**
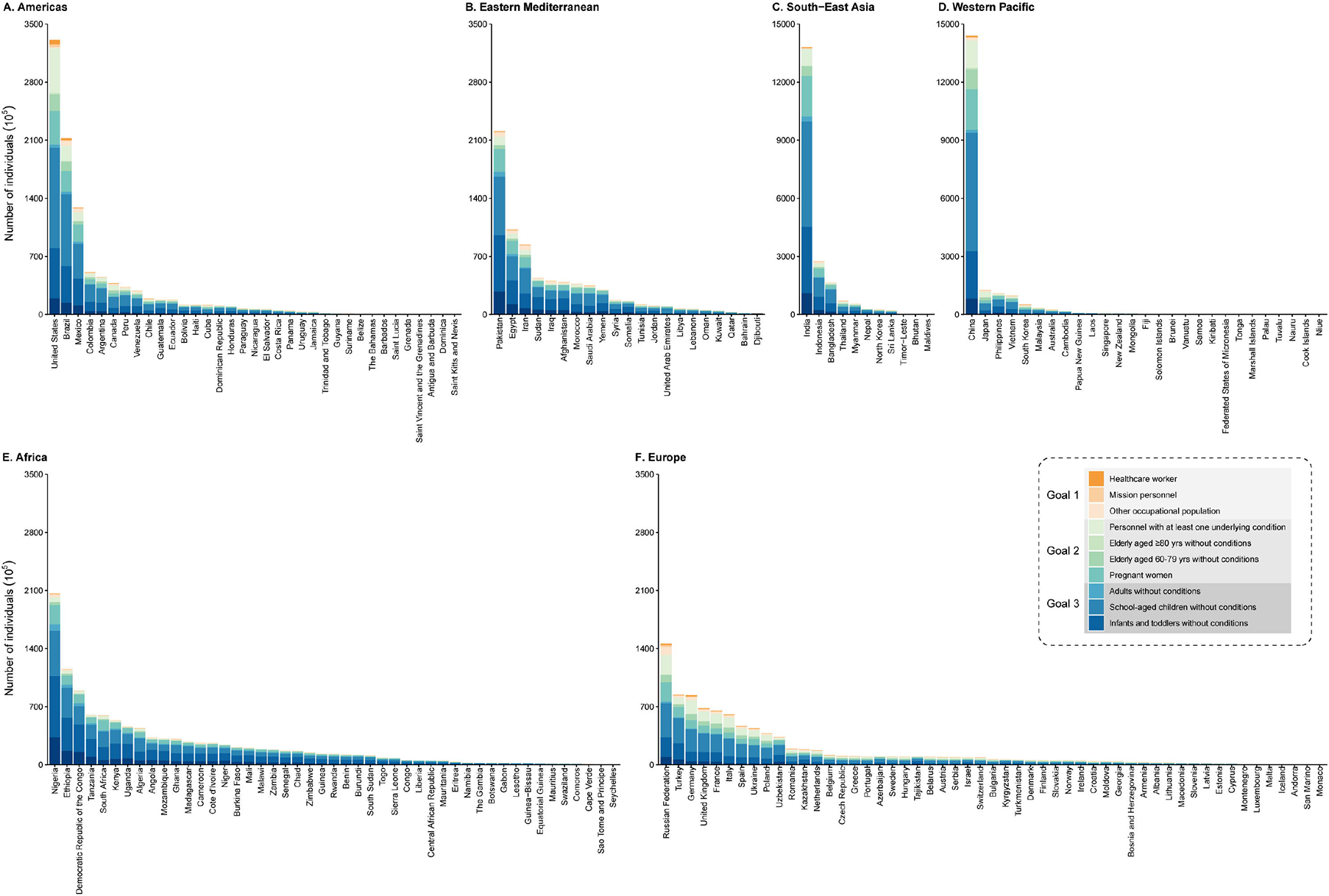
National estimates of the size of target population for COVID-19 vaccination, by goals of vaccination program.

**Figure 5.**
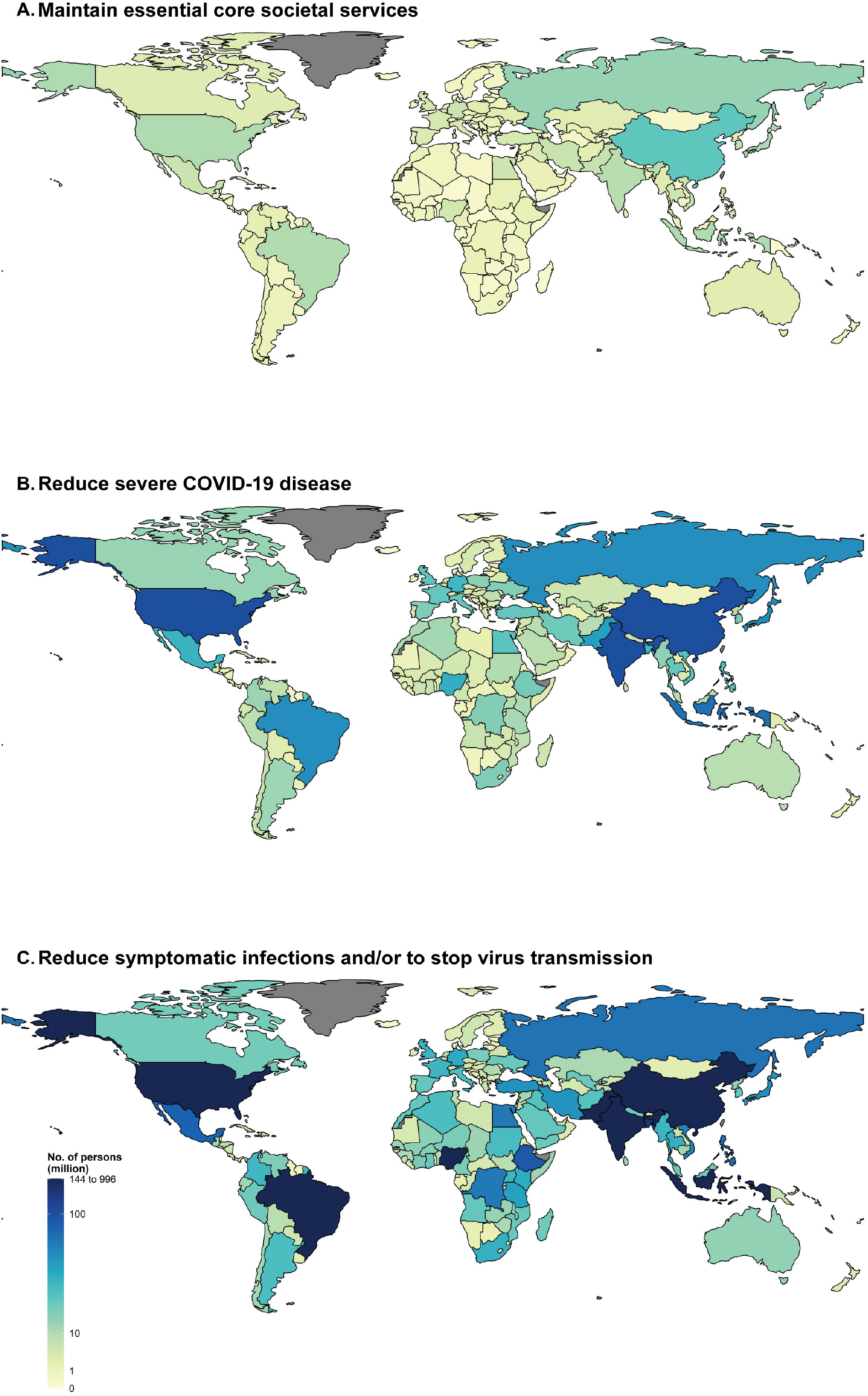
Estimates of the size of target populations for COVID-19 vaccination program across the globe, by goals of vaccination program. (A) target populations to maintain essential core societal services; (B) target populations to reduce severe COVID-19 disease; (C) target populations to reduce symptomatic infections and/or to stop virus transmission. Note that the empty refers to there is no population data available.

For the eleven countries lacking data on age-specific prevalence of underlying conditions, the obtained estimates of the target populations are robust to changes in assumptions about frequency of underlying conditions (compare Tab. S4 with Tab. S6).

## Discussion

We determined different target population groups according to 3 goals of the vaccination program (maintain essential societal functions, minimize severe disease, interrupt transmission), and quantified the size of each target group on a global, regional and country level. There are important variations in the amount of vaccines needed depending on the goals of the vaccination program, and the distribution of target populations varies within and between regions. In particular, large demand for vaccines was seen in essential workers and high-risk populations with poor health conditions, with the later accounting for 29.2% of total population.

The elderly with underlying conditions and occupational population with high-risk exposure are at higher risk and can benefit greatly from vaccination. However, the Expanded Program on Immunization proposed by WHO (*18*) currently targets infants and children, and countries have not yet expanded routine vaccination for the elderly, who have little access to the vaccine, especially in low- and middle-income countries. A representative survey showed that 67% of U.S. adult population would get a COVID-19 vaccine (*17*), indicating that vaccination acceptance still needs to be improved by minimizing vaccine hesitancy, and providing financial support and effective vaccination strategies that maximize health benefit through efficient use of limited resources. Given these limitations, between-country variations in vaccine acceptance and delivery need to be considered when tailoring COVID-19 vaccine allocation and distribution to each locale.

We estimated that approximately 15.6 billion doses of a COVID-19 vaccine will be requested by 194 WHO Member States for a universal COVID-19 vaccination program, given a two-dose regimen. Assuming all of the vaccine manufacturers with existing candidates can offer vaccines concurrently, the global production capacity of COVID-19 vaccine is estimated at 3.5 billion doses annually (*7*). Not enough vaccine will be available at the beginning of a COVID-19 vaccination program, even in an optimistic scenario. To achieve herd immunity by protecting at least 60-80% of individuals (4.7 to 6.2 billion), it will take about 17 to 22 months to produce enough COVID-19 vaccines. Vaccine supply and delivery services will constrain the roll-out of COVID-19 vaccination programs as well. In this context, the same barriers would apply to all target groups, but vaccination of targeted occupational or high-risk groups will likely be more feasible than the general public without any underlying conditions. This together comprise an estimated 2.51 billion people in 194 WHO Member States.

Within- and between-region disparities in the distribution of each target population highlight different demands for COVID-19 vaccine. These disparities will result in different durations of vaccination program, due to global limitations in vaccine production/supply capacity. For example, in countries with sufficient capacity for vaccine production and supply to meet the national demand (e.g., the United States and China), the COVID-19 vaccination program could last a few months, while it could last much longer in low- and middle- income countries which have relatively lower capacity for vaccine production and delivery (*18*). Thus, vaccine allocation plans need to be adjusted accordingly to consider inter- and intra-regional disparities in the demand for vaccine and capacity for vaccine production/supply. Besides direct benefits (i.e., protection from infection, reduction in illnesses and mortality rates), vaccine prioritization and allocation should also balance indirect benefits that can reduce virus circulation in a community, as vaccinated individuals are less likely to be infected and transmit the virus (*19*). In particular, indirect benefits may be important to protect individuals aged >65 years of age who are at increased risk of severe disease and also possibly less likely to be directly protected by vaccination due to immune senescence (*20*).

We were unable to collect data on population stratified by occupation in 25.8% of the countries considered. Excluding those from the analysis would lead to the exclusion of a substantial proportion of the total population (∼0.86 billion people, 11%), and would make it difficult to understand global heterogeneity in the distribution of each target population. Therefore, in this study, missing data were tackled by MICE algorithm (*16*) in which biases can be overcome and incomplete data are allowed to be included in analyses. Unlike other approaches using values imputed from average numbers, however, MCIE algorithm allows for the uncertainty about the missing values by creating several different plausible imputed datasets from their predictive distribution (based on the observed data). Variability between the imputed datasets can be considered, and average estimates can be obtained (*21*). This gives more robust estimates of the size of target populations on local, regional, and global scales.

A few limitations should be highlighted in this study. Lack of timely data for 2020 constrains estimates of population sizes in many countries. However, the distribution of target occupational and high-risk groups is likely stable over a few years. Second, we could not explore within-country variations in target populations. Actual vaccine allocation plans should be carefully investigated in relation to the policy decisions of each population when relevant data are available in a given country. Third, due to data availability, we cannot provide estimates for the size of target population by other demographic factors, such as racial and ethnic groups which are reported to be risk factors for COVID-19 risk and adverse outcomes (*22, 23*). In addition, given relatively lower prevalence of underlying conditions among essential workers aged <60 years of age (5.8%, 95%CI 5.7-6.1%) (*24*), we did not subtract these essential workers with underlying conditions from the broader group of adults with underlying conditions.

In conclusion, findings from this study provide evidence base for global, regional, and national vaccine prioritization and allocation plan. Within and between-region variations in the size of target populations emphasize the tenuous balance between vaccine demand and supply, especially in low- and middle-income countries without sufficient capacity to meet domestic demand for COVID-19 vaccine. Moreover, in a given country, vaccine prioritization and allocation should be targeted towards on the basis of specific health or societal objectives, and local variations at the individual or regional levels.

## Data Availability

The datasets used and analyzed during the current study are available in appendix.

## Contributors

W. Wang, Q. Wu, and H. Yu had full access to all of the data in the study and take responsibility for the integrity of the data and the accuracy of the data analysis. C. Viboud, M. Ajelli, and H. Yu were responsible for its conception and design. W. Wang, Q. Wu, K. Dong, X. Chen, X. Bai, X. Chen, and Z. Chen were responsible for the acquisition, analysis, or interpretation of data. W. Wang, Q. Wu, J. Yang, C. Viboud, M. Ajelli, and H. Yu drafted the manuscript. J. Yang, C. Viboud, M. Ajelli, H. Yu made critical revision of the manuscript for important intellectual content. W. Wang, Q. Wu, X. Chen, and X. Bai did the data analysis. J. Yang, C. Viboud and M. Ajelli provided administrative, technical, or material support.

## Acknowledgments

The study was funded by the National Science Fund for Distinguished Young Scholars (No. 81525023) and National Science and Technology Major Project of China (No. 2018ZX10713001-007, No. 2017ZX10103009-005, No. 2018ZX10201001-010).

---

**Table 1.** Estimates of target population sizes for COVID-19 vaccination, by goals of vaccination program.

|  | All | WHO region (million) |  |  |  |  |  |
| --- | --- | --- | --- | --- | --- | --- | --- |
|  |  | Africa | Americas | Eastern Mediterranean | Europe | Western Pacific | South-East Asia |
| Maintaining essential core societal services |  |  |  |  |  |  |  |
| Subtotal | 246.9 | 19.0 | 41.8 | 31.6 | 63.8 | 58.8 | 32.0 |
| Healthcare workers | 40.7 | 1.4 | 10.8 | 1.8 | 10.8 | 10.6 | 5.2 |
| Mission personnel<sup>a</sup> | 46.9 | 3.2 | 8.0 | 6.1 | 8.4 | 10.7 | 10.4 |
| Other occupational population | 159.4 | 14.4 | 23.0 | 23.7 | 44.6 | 37.4 | 16.3 |
| Reducing severe COVID-19 disease |  |  |  |  |  |  |  |
| Subtotal | 2260.4 | 232.1 | 311.5 | 173.2 | 352.4 | 642.5 | 548.8 |
| Elderly aged ≥60 years-old with at least one underlying conditions | 657.6 | 35.8 | 113.1 | 35.9 | 144.8 | 197.9 | 130.2 |
| Elderly aged ≥80 years-old without any underlying conditions | 28.6 | 0.8 | 4.5 | 0.8 | 7.3 | 10.7 | 4.4 |
| Elderly aged 60-79 years-old without any underlying conditions | 354.1 | 19.0 | 51.2 | 15.8 | 61.5 | 132.0 | 74.6 |
| Adults aged <60 years-old with at least one underlying conditions | 1078.1 | 137.5 | 127.7 | 102.0 | 127.8 | 279.1 | 304.0 |
| Pregnant women | 142.0 | 39.1 | 14.9 | 18.7 | 11.0 | 22.7 | 35.6 |
| Reducing symptomatic infections and/or to stop virus transmission |  |  |  |  |  |  |  |
| Subtotal | 5242.7 | 869.0 | 664.8 | 521.0 | 516.7 | 1230.5 | 1440.7 |
| Adults without conditions | 2772.4 | 310.6 | 375.7 | 231.9 | 302.2 | 771.6 | 780.4 |
| School-age individuals without conditions | 1816.1 | 390.2 | 217.4 | 206.3 | 159.6 | 343.8 | 498.9 |
| Infants and toddlers without conditions | 654.2 | 168.2 | 71.8 | 82.8 | 54.9 | 115.2 | 161.3 |
| Total | 7750.0 | 1120.2 | 1018.1 | 725.7 | 932.9 | 1931.7 | 2021.4 |

<sup>a</sup>Estimated number of individuals is based on 149 countries with available data.

